# Negative Excess Mortality in Pneumonia Death caused by COVID-19 in Japan

**DOI:** 10.1101/2021.01.22.21250283

**Authors:** Junko Kurita, Tamie Sugawara, Yoshiyuki Sugishita, Yasushi Ohkusa

## Abstract

**Background:** Since the emergence of COVID-19, cases of excess mortality from all causes have been very few in Japan.

**Object:** To evaluate COVID-19 effects precisely, we specifically examine deaths caused by pneumonia and examine excess mortality attributable to pneumonia in Japan.

**Method:** We applied the NIID model to pneumonia deaths from 2005 up through November, 2020 for the whole of Japan. Introduction of routine pneumococcal vaccination for elderly people and revision in ICD10 were incorporated into the estimation model.

**Results:** No excess mortality was found for 2020. However, negative excess mortality was observed as 178 in May, 314 in June, and 75 in July. No negative excess mortality was not found between August and November.

**Discussion and Conclusion:** Significantly negative excess mortality might reflect precautions taken by people including wearing masks, washing hands with alcohol, and maintaining social distance. They reduced the infection risk not only of for COVID-19 but also of other infectious diseases causing pneumonia.

## 1. Introduction

Since the emergence of COVID-19, excess mortality from all causes has been low in Japan [1]. Actually, data obtained throughout Japan show 12 and 104 cases of excess mortality in August and October, 2020. These very few cases of excess mortality in Japan might be attributable to the lower number of traffic accidents from voluntary restrictions against going out. Therefore, we specifically examine deaths caused by pneumonia for precise evaluation of COVID-19 effects on Japan. The present study examines excess mortality attributable to pneumonia in Japan in 2020.

Regarding pneumonia deaths, two points can be ignored among all causes of death. The first point is pneumococcal vaccine for elderly people. In Japan, routine immunization was introduced in October, 2015. We can expect continuous declines in pneumonia deaths.

The second point is a change in International Classification of Diseases ver. 10 (ICD10) from the 2003 version to the 2013 version. In Japan, the switch occurred in 2018. The number of pneumonia deaths was expected to be reduced by 20% [2]. Therefore, those points are incorporated into the model for excess mortality related to pneumonia deaths.

## 2. Method

The procedure of estimation was almost identical to that used for an earlier study [1], except for some points. Excess mortality was defined as the difference between the actual number of deaths and an epidemiological threshold if the actual number of deaths was larger than an epidemiological threshold. The epidemiological threshold is defined as the upper bound of the 95% confidence interval (CI) of the baseline. The baseline is defined as the number of deaths which are likely to have occurred if an influenza outbreak had not occurred. Therefore, if the actual deaths were fewer than the epidemiological threshold, then excess mortality was not inferred. Also, we defined negative excess mortality as the difference between the actual number of deaths and the lower bound of 95% CI if the actual number of deaths was less than the lower bound of 95% CI.

The data used for this study were monthly deaths of all causes reported from 2005 through November 2020 [3]. The NIID model, the Stochastic Frontier Estimation [4–10], is presented as

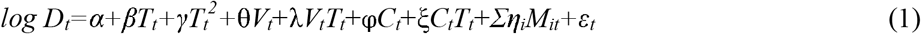

and

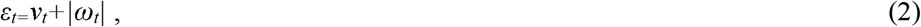

where *D*_*t*_ represents pneumonia deaths in month/year *t, T*_*t*_ denotes the linear time trend, *V*_*t*_ is a dummy variable for routine pneumococcal vaccination, *C*_*t*_ is a dummy variable for revision in ICD10, and *M*_*it*_ is a dummy variable for the month. That is, *V*_*t*_ is one after October, 2014 and otherwise zero. *C*_*t*_ is one after 2018 and otherwise zero. *M*_*it*_ is one if *t* is the *i-*th month and otherwise zero. Moreover, *ν*_*t*_ and *ω*_*t*_ are stochastic variables as *ν*_*t*_*∼N*(*0, μ*^2^) and *ω*_*t*_ *∼N*(*0,ξ*^2^); they are mutually independent. Although *ν*_*t*_ represents stochastic disturbances, *ω*_*t*_ denotes non-negative deaths attributable to influenza. These disturbance terms in this model are parameterized by two parameters: *ξ*/*μ* and (*μ*^*2*^*+ξ*^*2*^)^*0*.*5*^. If the null hypothesis *ξ*/*μ*=0 is not rejected, then the Stochastic Frontier Estimation model is inappropriate.

The study area was the entire nation of Japan. The study period for estimation was 2005 through November 2020. We adopted 5% as the level at which significance was inferred for results. Pneumonia is defined as J12–J18 in ICD10.

## 3. Results

Because the estimated coefficients of the cross term of routine vaccination initiation and time trend, the ICD10 revision dummy, and the cross term of ICD10 revision and time trend were not found to be significant, we dropped these terms from the estimation equation (1). Table 1 presents estimation results without these variables. The estimated coefficients of ICD10 revision and its cross term with the time trend were not significant. Moreover, although the estimated coefficients of routine pneumococcal vaccination for elderly were significant, its cross term with a time trend was not significant.

**Table 1.** NIID Model estimation results for pneumonia death from 2005 through November 2020 in Japan

| Explanatory variable | Estimated coefficient | *p*-value |
| --- | --- | --- |
| Constant | 9.0377 | < 0.0004 |
| Time trend | 0.006438 | < 0.0004 |
| Time trend <sup>2</sup> | -0.0000162 | < 0.0004 |
| Vaccination | 0.1279 | < 0.0004 |
| January | 0.1233 | < 0.0004 |
| February | -0.02197 | 0.189 |
| March | -0.01030 | 0.303 |
| April | -0.1029 | < 0.0004 |
| May | -0.1352 | < 0.0004 |
| June | -0.2846 | < 0.0004 |
| July | -0.2653 | < 0.0004 |
| August | -0.2257 | < 0.0004 |
| September | -0.2548 | < 0.0004 |
| October | -0.1749 | < 0.0004 |
| November | -0.1028 | < 0.0004 |
| $\xi/\mu$ | 2.085 | < 0.0004 |
| $(\mu^2 + \xi^2)^{0.5}$ | 0.08992 | < 0.0004 |

Note: The observations were 188. The log likelihood was 319.594. $\xi^2$ denotes the variance of the non-negative disturbance term. $\mu^2$ represents the variance of the disturbance term. “Vaccination” is a dummy variable for routine pneumococcal vaccination: one after October 2014 and zero otherwise.

Figure 1 presents observed deaths, the estimated baseline, and its threshold based on the model including insignificant terms. Figure 2 specifically depicts the last 12 months in Japan. We found no excess mortality in 2020. In fact, we observed negative excess mortality as 178 in May, 314 in June, and 75 in July. No negative excess mortality was not found between August and November.

**Figure 1:**
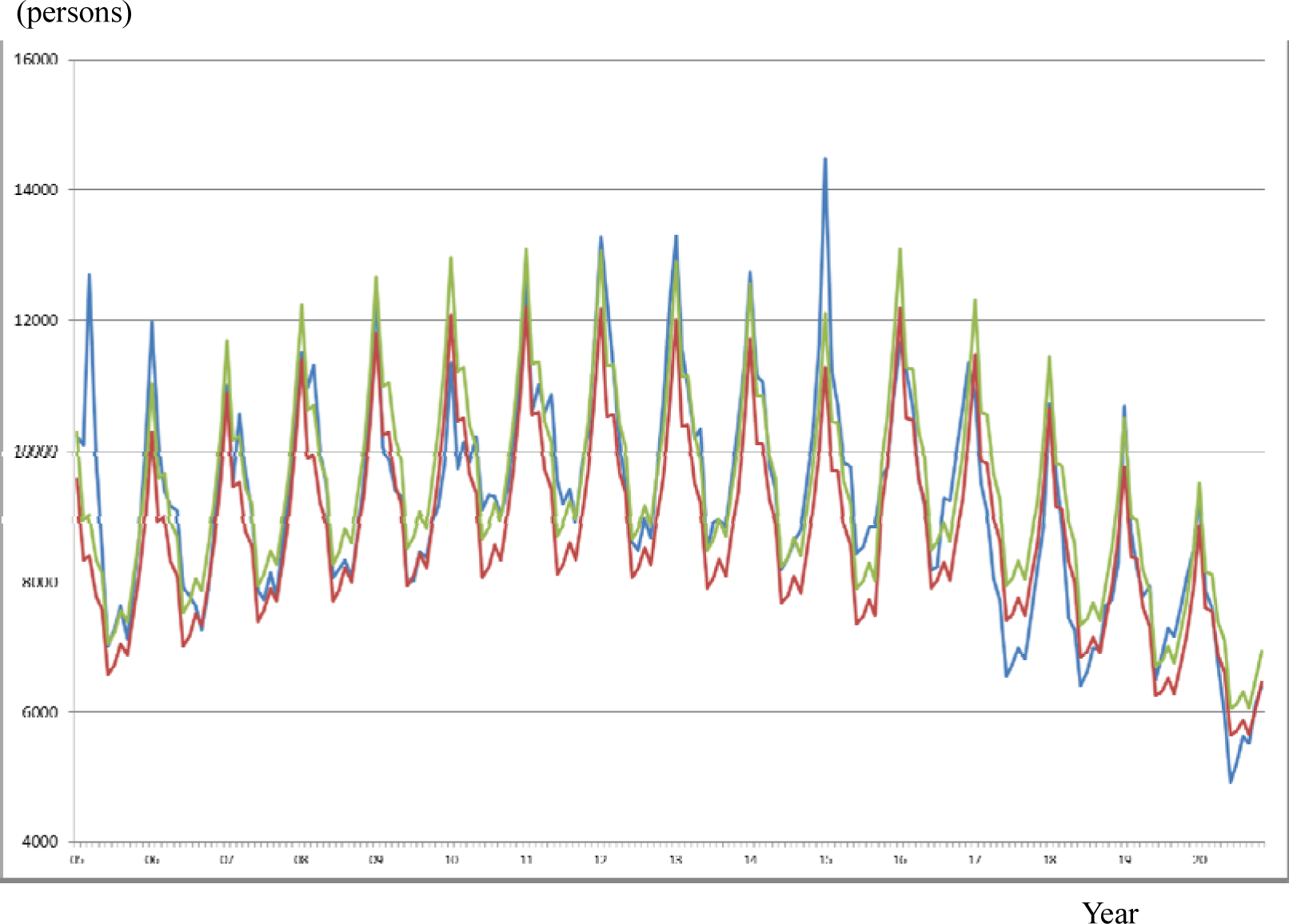
Observations of the estimated baseline and threshold for pneumonia deaths from 2005 through November 2020 in Japan. Note: The blue line represents observations. The orange line represents the estimated baseline. The gray line shows its threshold.

**Figure 2:**
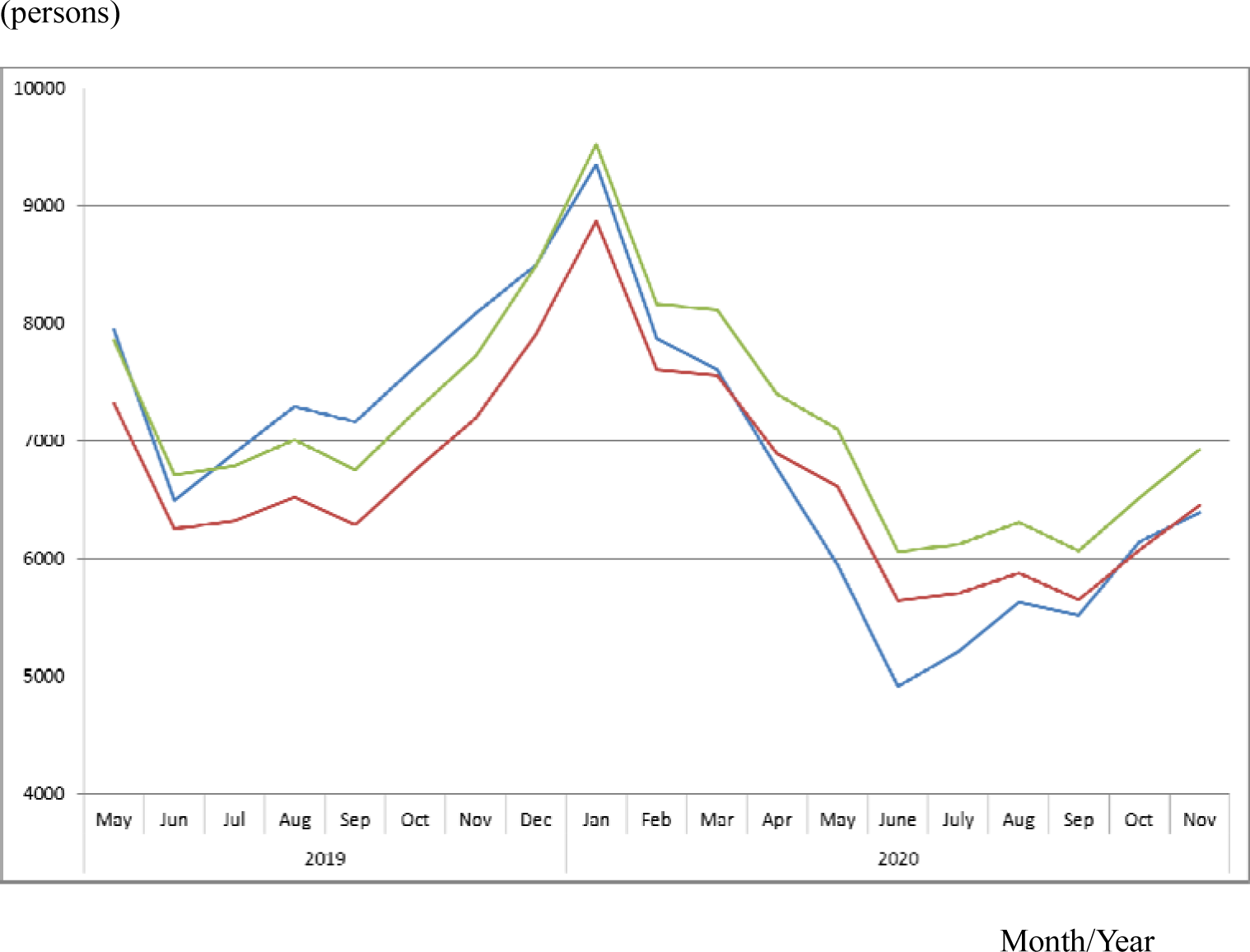
Observations of the estimated baseline and threshold for pneumonia deaths since May 2019 in Japan. Note: The blue line represents observations. The orange line represents the estimated baseline. The gray line shows its threshold.

## 4. Discussion

This study applied the NIID model to pneumonia deaths to detect excess mortality attributable to COVID-19. No excess mortality was found in 2020, but 1074 negative excess mortality cases were identified in the last three months. Its volume was about 3.1% of the baseline in the corresponding period.

Throughout Japan, about 4000 cases of mortality caused by COVID-19, as confirmed by PCR testing, were reported to the MHLW officially as of December [12]. Therefore, even if COVID-19 actually caused external mortality, neither the NIID model nor another statistical model would detect significant effects attributable to COVID-19. Of course, it is possible that some deaths caused by COVID-19 had been classified as pneumonia deaths.

Even so, COVID-19 is associated with significantly reduced pneumonia mortality in May, June, and July, in Japan. The result was not attributable to the wide spread of pneumococcal vaccination or revision in ICD10 because some excess mortality was found in 2019. Results suggest that precautions among people, including wearing masks, washing hands with alcohol, and maintaining social distance reduced the infection risk not only of COVID-19 but of other infectious diseases causing pneumonia. Therefore, pneumonia deaths decreased, completely offsetting the COVID-19 effects.

Some researchers in Japan have emphasized considerable excess mortality from all causes of death through October of around 19 thousand at maximum due to COVID-19 [11] when using the Farrington algorithm [12] and EuroMOMO [13], which was 11 times larger than the number of death confirmed by PCR test. This study measured excess mortalities as the gap between observation and beeline, not threshold as, in prefectures where observation was higher than threshold. Therefore, their estimated too huge excess mortality may seriously mislead the risk participation for COVID-19 among the general population.

The effect of ICD10 revision was not significant at all. It is probably attributable to overlapping of its period with routine pneumococcal vaccination. In other words, its effect was not so great if one incorporates vaccination effects into the estimation model. Moreover, the cross-term of vaccination with the time trend was not significant, although Figure 1 presents a clear decrease in the number of pneumonia deaths. That
result is probably attributable to the significant negative square term of the time trend. Actually, before 2015, the increasing pace was decreasing gradually and almost flat in 2013 or 2014. It might be reflected by the gradual spread of peumoccocal vaccine before routine immunization was introduced. Therefore, the negative time trend squared might be explained already by the decrease since 2015.

The present study has some limitations. First, our results reflected data through November, 2020 when the outbreak of COVID-19 was not severe. Subsequently, and especially in winter, a severer COVID-19 outbreak emerged. Negative excess mortality from pneumonia deaths might disappear. Continuous monitoring is expected to be necessary.

Second, by rules governing the determination of the cause of death, some cases of death attributable to COVID-19 were not classified as pneumonia or as COVID-19 infection. The rule for cause of death requires classification by the most fundamental cause. For example, if cancer patients were infected by COVID-19, but then showed pneumonia and died, their proximate cause of death was probably cancer, and not COVID-19 or pneumonia. Therefore, some fraction of negative excess mortality by pneumonia might be classified as caused by other than pneumonia. However, even for periods when deaths of all causes in some months were fewer than the baseline, but not significantly in 2020 [1], the total deaths during the entire COVID-19 outbreak period were fewer than the usual level.

Thirdly, similarly to the point raised earlier, if a healthy person was presumed to be infected with COVID-19, then showed pneumonia, and finally died, then the cause of death should be classified as COVID-19, or as U07.1, for an identified virus or as U07.12 for an unidentified virus under ICD10. Conversely, for a case in which a healthy person tested positive for COVID-19, showed pneumonia, and died, but who was not presumed to be infected by COVID-19, the cause of death should be classified as pneumonia. Therefore, negative excess mortality related to pneumonia does not necessarily represent negative excess mortality of COVID-19 itself.

## 5. Conclusion

We found no excess mortality in 2020, but 1074 cases of negative excess mortality between May and July. Continued careful monitoring of excess mortality of COVID-19 is expected to be important.

The present study is based on the authors’ opinions: it does not reflect any stance or policy of their professionally affiliated bodies.

## Data Availability

Ministry of Health, Labour and Welfare

## 6. Acknowledgement

We acknowledge Dr. Nobuhiko Okabe, Kawasaki City Institute for Public Health, Dr.Kiyosu Taniguchi, National Hospital Organization Mie National Hospital, and Dr.Nahoko Shindo, WHO for their helpful support.

## 8. Ethical considerations

All information used for this study was published on the web site of MHLW [12]. Therefore, no ethical issue is presented.

